# Quantifying and mitigating the impact of the COVID-19 pandemic on outcomes in colorectal cancer

**DOI:** 10.1101/2020.04.28.20083170

**Authors:** Amit Sud, Michael Jones, John Broggio, Stephen Scott, Chey Loveday, Bethany Torr, Alice Garrett, David L. Nicol, Shaman Jhanji, Stephen A. Boyce, Matthew Williams, Georgios Lyratzopoulos, Claire Barry, Elio Riboli, Emma Kipps, Ethna McFerran, Mark Lawler, David C. Muller, Muti Abulafi, Richard Houlston, Clare Turnbull

**Affiliations:** Division of Genetics and Epidemiology, Institute of Cancer Research, London, UK; National Cancer Registration and Analysis Service, Public Health England, Wellington House, London, UK; RM Partners, West London Cancer Alliance, London, UK; Urology Unit, Royal Marsden NHS Foundation Trust, London, UK; Department of Clinical Studies, Institute of Cancer Research, London, UK; Department of Anaesthesia, Perioperative Medicine and Critical Care, Royal Marsden NHS Foundation Trust, London, UK; Division of Cancer Biology, Institute of Cancer Research, London, UK; Department of Colorectal Surgery, Oxford University Hospitals NHS Foundation Trust, Oxford, UK; Department of Clinical Oncology, Imperial College Healthcare NHS Trust, London, UK; Computational Oncology Group, Imperial College London, London, UK; Epidemiology of Cancer Healthcare and Outcomes (ECHO) Group, University College London, London, UK; School of Public Health, Imperial College London; The Breast Unit, Royal Marsden NHS Foundation Trust, London, UK; Patrick G Johnston Centre for Cancer Research, School of Medicine, Dentistry and Biomedical Sciences, Queen's University Belfast, Belfast; Colorectal Surgery, Croydon Health Services NHS Trust, on behalf of RMP NICE FIT Steering Group; Department of Clinical Genetics, Royal Marsden NHS Foundation Trust, London, UK

**Author notes:** These authors contributed equally to the work (Amit Sud, Michael Jones, John Broggio, Stephen Scott). Corresponding author: Clare Turnbull; address: Division of Genetics and Epidemiology, Institute of Cancer Research, London, UK, SM2 5NG.

## Abstract

**Background:** The COVID-19 pandemic has caused disruption across cancer pathways for diagnosis and treatment. In England, 32% of colorectal cancer (CRC) is diagnosed via urgent symptomatic referral from primary care, the “2-week-wait” (2WW) pathway. Access to routine endoscopy is likely to be a critical bottleneck causing delays in CRC management due to chronic limitation in capacity, acute competition for physician time, and safety concerns.

**Methods:** We used age-specific, stage-specific 10 year CRC survival for England 2007–2017 and 2WW CRC cases volumes. We used per-day hazard ratios of CRC survival generated from observational studies of CRC diagnosis-to-treatment interval to model the effect of different durations of per-patient delay. We utilised data from a large London observational study of faecal immunochemical testing (FIT) in symptomatic patients to model FIT-triage to mitigate delay to colonoscopy.

**Findings:** Modest delays result in significant reduction in survival from CRC with a 4-month delay resulting across age groups in ≥20% reduction in survival in Stage 3 disease and in total over a year, 1,419 attributable deaths across the 11,266 CRC patients diagnosed via the 2WW pathway. FIT triage of >10 ug Hb/g would salvage 1,292/1,419 of the attributable deaths and reduce colonoscopy requirements by >80%. Diagnostic colonoscopy offers net survival in all age groups, providing nosocomial COVID-19 infection rates are kept low (<2·5%).

**Interpretation:** To avoid significant numbers of avoidable deaths from CRC, normal diagnostic and surgical throughput must be maintained. An accrued backlog of cases will present to primary care following release of lockdown, supranormal endoscopy capacity will be required to manage this without undue delays. FIT-triage of symptomatic cases provides a rational approach by which to avoid patient delay and mitigate pressure on capacity in endoscopy. This would also reduce exposure to nosocomial COVID-19 infection, relevant in particular to older patient groups.

**Funding:** Breast Cancer Now, Cancer Research UK, Bobby Moore Fund for Cancer Research, National Institute for Health Research (NIHR).

## INTRODUCTION

The COVID-19 pandemic has placed unprecedented pressure on healthcare services. Urgent redeployment of staff towards the management of COVID-19 cases within primary and secondary care has required deprioritisation of non-COVID-19-related non-emergency clinical services. For many conditions, delay to treatment will impact the quality of life but is unlikely to have long-term consequences. For patients with localised cancer, delay to diagnosis and treatment will increase the likelihood of metastatic disease, with some patients’ tumours progressing from being curable by surgery (or radiotherapy) with near-normal life expectancy to being incurable, with very limited life expectancy^1^. The “two-week wait” (2WW) referral protocols were established to ensure a rapid access pathway to treatment for patients presenting with classic ‘red-flag’ symptoms suggestive of a particular cancer type. Lockdown, public anxiety, and disruption of primary care services have all contributed to the significant reduction in presentation and referral of symptomatic patients from primary into secondary care^2^. For symptomatic patients reaching primary care, the high mortality from COVID infection in older patients in some areas, raised concern regarding the risk-benefit trade-off which has hampered hospital referral^3^. There is likely to be a contraction in healthcare capacity for non-COVID-19 care until full resolution of the pandemic, with wide ongoing disruption.

Colorectal cancer (CRC) is the fourth most common cancer and second most common cause of cancer deaths in the UK^4^. CRC will likely be particularly impacted by the COVID-19-related disruption at many points in the pathway to treatment. Laparoscopic resections have been discontinued on account of risk of aerosol generation; open operations require sizeable inpatient stay and on occasion intensive care (ICU)^5^. Shutdown due to safety concerns and redeployment of consultant physicians has dramatically constricted availability of endoscopy^6^. There has been effective discontinuation of the National Bowel Cancer Screening program, which normally accounts for diagnosis of 10% of CRC, most of which are at early stage^7^. Normally, around 50% of CRC is diagnosed at stage 1 or 2, and over a quarter of cases present as emergencies^8–10^. Delay to diagnosis as a result of the COVID-related healthcare disruption will lead to upstaging of CRC in many patients, inevitably with worse prognosis. Although not previously standard-of-care, since late March 2020 in some regions of the UK, faecal immunochemical testing (FIT) has been implemented as a strategy to triage symptomatic patients to mitigate the reduced access to endoscopy^11,12^.

To inform decision-making we examined the impact of varying putative periods of delay to diagnosis/surgical management to CRC outcome in urgent symptomatic 2WW patients, accounting for COVID-19-associated mortality during hospitalisation. We also examined the impact of implementing FIT-triage of symptomatic patients and considered the per-patient risk/benefit profile for diagnostic colonoscopy at different rates of nosocomial infection.

## METHODS

### Data sources

We obtained five (2013–2017) and 10-year (2008–2017) survival from CRC from Public Health England National Cancer Registration Service (NCRAS)^9^. We obtained data on route to diagnosis with distribution by age and stage for CRC from NHS England Clinical Commissioning Groups^8^. For conversion rates from referrals for suspected cancer to cancer diagnoses, we used data from Cancer Waits/Faster Diagnosis Standard data for West London 2019/20^13^. Median duration of hospital stay was based on information from three large UK surgical oncology centres. We used data from Wuhan as the basis for mortality associated with COVID-19, as no age-specific UK mortality data is currently available except for ICU patients^3,14^. Actuarial survival was based on UK Office of National Statistics life tables for 2016–2018^15^. We obtained data on positive predictive value (PPV), negative predictive value (NPV), sensitivity, and positivity for FIT at different thresholds from the UK NICE FIT study, in which 9,822 symptomatic patients received a FIT-assay before colonoscopy^16,17^.

### Analysis

#### Impact of COVID-associated delay on outcomes

Given statistical cure rates in CRC patients are known to occur 7–8 years post-diagnosis, we used 10-year stage-specific survival data in our calculations. Because no such data were directly available for recently diagnosed cohorts, we have estimated these by applying the ratio of stage-specific/all-stage 5-year survival data to 10-year all-stage data^9,18^. Net survival estimates were used whereby background age-specific death rates have been adjusted to reflect cancer-specific mortality. To estimate per day hazard ratios (HRs) for mortality, we used published data on overall survival (OS) from CRC for different diagnosis-to-treatment intervals (DTI), which therefore includes within-stage progression, between-stage progression, and emergency presentation occurring within the interval^19^. To estimate 10-year survival under current conditions, we applied the 10-year survival under standard care and adjusted for COVID-related post-surgical mortality. To estimate 10-year survival following delay to surgery, we applied to standard 10-year survival the HR relating to the specified number of days of delay and included COVID-related post-surgical mortality.

We estimated COVID-associated surgical mortality based on per day rate of nosocomial infection, operation-specific duration of post-surgical admission and age-specific mortality from infection. Assuming ongoing evolution in cold protocols, segregation, and testing of staff and patients, we modelled rates of nosocomial COVID infection would halve every three months. Where not under evaluation, the nosocomial infection rate was assumed to be 5% per day. We used actuarial life-expectancies, to estimate life years gained, averaged per patient.

We considered per-patient delay of up to 6-months considering a 1-year period of disruption, examining the reduction in survival and life years gained (LYG), comparing by age and stage surgery under standard care, current conditions and post-delay.

We quantified the numbers of cancers diagnosed by age and stage through the urgent symptomatic 2WW pathway, estimating referral numbers from the conversion rates from referrals for suspected cancer to cancer diagnoses.

#### Impact of implementing FIT to triage symptomatic patients

We examined adopting FIT thresholds to identify a subset of symptomatic referrals for prompt investigation and management. We examined FIT thresholds of 2, 10, and 150 ug Hb/g faeces as these are the thresholds utilised in the NICE FIT study and are currently being piloted^12^. We assumed FIT-positive cases would be managed without delay as per standard care, but that FIT-negative CRC cases would only be diagnosed following delayed colonoscopy. For the different FIT thresholds, for each age-/stage-specific stratum, we calculated the sum of deaths and lost life years mitigated, with associated colonoscopy requirements.

#### Impact of COVID-associated risk on risk/benefit of colonoscopy and surgery

The likelihood of COVID infection resulting from colonoscopy was calculated by applying varying rates of nosocomial infection risk (estimated as being a half-day of risk per day rate of nosocomial infection). We combined this with the age-specific mortality if infected^3^, to estimate COVID-related mortality associated with colonoscopy. We factored a “technical” mortality risk from perforation of 1 in 10,000 and combined the two to produce a combined per-procedure mortality^20^.

Based on the conversion rate from referral to diagnosis of CRC, we calculated for each age-stage stratum the survival benefit per patient undergoing colonoscopy. We considered opportunity for delay of 2, 4, and 6 months and rates of nosocomial infection per procedure of 5% (very high), 2·5% (high), and 1% (moderate), assuming that nosocomial rate would halve every 3 months through improvements in practice. Under each scenario, to assess risk-benefit by age-group of diagnostic colonoscopy versus delay, we compared the CRC survival benefit against the mortality risk (COVID-19 and technical). We performed equivalent analysis for patients diagnosed with CRC, comparing nosocomial infection with survival benefit from surgery.

## RESULTS

Delays in diagnosis and management were found to be associated with substantial decrements in the 10-year survival of CRC patients, though the impact varied notably by age, period of delay, and tumour stage. For stage 3 CRC across all ages, a two month delay to surgery is predicted to cause >9% reduction in survival and for a six month delay >29%. Even for stage 1 CRC, the effect of delaying surgery is considerable; a 4 month delay for Stage 1 disease is likely to result in a 6·8% reduction in survival for those aged 70–79 and 17·2% in those over 80 (**Table 1**). Considering the impact on actuarial life years lost, the impact of delayed management of CRC is particularly high for patients with either stage 2 or stage 3 cancers. For example, a 4-month delay in surgery for a stage 3 CRC patient will lead to 9·4 life years lost on average to patients aged 30–39 and 15·1 life years lost for 6 month delay (**Supplementary Table 1**). In England in 2013–2016, on average 32% (11,229/34,863) of CRC diagnosed came through urgent (2WW) referral; an average delay of 4 months over a single year would conservatively result in 1,419 deaths and 20,315 lost life years by this route to CRC diagnosis alone (**Table 2, Table 3**).

**Table 1:** Reduction in 10-year survival from colorectal cancer from delay to surgery (Assumes per day rate of COVID-19 nosocomial infection of 5% at T_0_ and 1·25% after 6 months).

|  |  | 2 months | 4 months | 6 months |
| --- | --- | --- | --- | --- |
| 30-39 years | Stage 1 | 0.4% | 1.1% | 1.9% |
|  | Stage 2 | 5.1% | 11.7% | 20.1% |
|  | Stage 3 | 9.1% | 20.0% | 32.2% |
| 40-49 years | Stage 1 | 1.6% | 3.9% | 7.0% |
|  | Stage 2 | 5.2% | 12.0% | 20.7% |
|  | Stage 3 | 9.7% | 21.1% | 33.5% |
| 50-59 years | Stage 1 | 1.7% | 4.1% | 7.5% |
|  | Stage 2 | 4.9% | 11.3% | 19.5% |
|  | Stage 3 | 9.3% | 20.5% | 32.8% |
| 60-69 years | Stage 1 | 1.7% | 4.3% | 8.0% |
|  | Stage 2 | 5.3% | 12.4% | 21.4% |
|  | Stage 3 | 9.5% | 20.8% | 33.1% |
| 70-79 years | Stage 1 | 2.7% | 6.8% | 12.5% |
|  | Stage 2 | 6.5% | 15.0% | 25.5% |
|  | Stage 3 | 11.0% | 23.2% | 35.0% |
| 80+ years | Stage 1 | 7.5% | 17.2% | 28.7% |
|  | Stage 2 | 8.2% | 18.5% | 30.4% |
|  | Stage 3 | 11.5% | 22.0% | 29.7% |

**Table 2:**
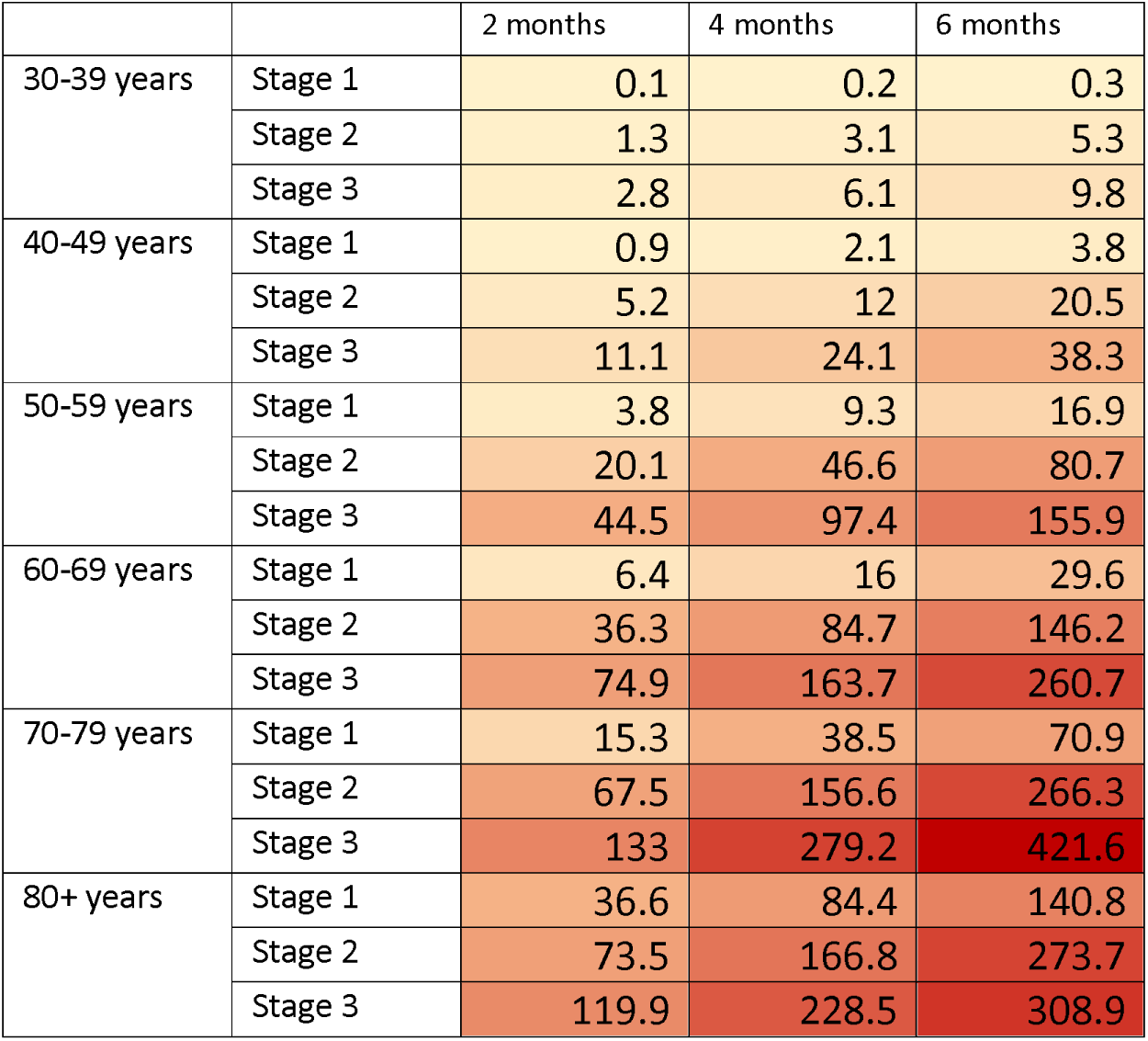
Attributable deaths over one year from delays for patients diagnosed with CRC via the 2-week-wait route to diagnosis (Assumes per day rate of COVID-19 nosocomial infection of 5% at T_0_ and 1·25% after 6 months).

|  |  | 2 months | 4 months | 6 months |
| --- | --- | --- | --- | --- |
| 30-39 years | Stage 1 | 0.1 | 0.2 | 0.3 |
|  | Stage 2 | 1.3 | 3.1 | 5.3 |
|  | Stage 3 | 2.8 | 6.1 | 9.8 |
| 40-49 years | Stage 1 | 0.9 | 2.1 | 3.8 |
|  | Stage 2 | 5.2 | 12 | 20.5 |
|  | Stage 3 | 11.1 | 24.1 | 38.3 |
| 50-59 years | Stage 1 | 3.8 | 9.3 | 16.9 |
|  | Stage 2 | 20.1 | 46.6 | 80.7 |
|  | Stage 3 | 44.5 | 97.4 | 155.9 |
| 60-69 years | Stage 1 | 6.4 | 16 | 29.6 |
|  | Stage 2 | 36.3 | 84.7 | 146.2 |
|  | Stage 3 | 74.9 | 163.7 | 260.7 |
| 70-79 years | Stage 1 | 15.3 | 38.5 | 70.9 |
|  | Stage 2 | 67.5 | 156.6 | 266.3 |
|  | Stage 3 | 133 | 279.2 | 421.6 |
| 80+ years | Stage 1 | 36.6 | 84.4 | 140.8 |
|  | Stage 2 | 73.5 | 166.8 | 273.7 |
|  | Stage 3 | 119.9 | 228.5 | 308.9 |

**Table 3:** Impact of 2, 4, and 6 month delay for colorectal cancer treatment and impact of salvage using FIT triage (Assumes per day rate of COVID-19 nosocomial infection of 5% at T_0_ and 1·25% after 6 months).

| Reference period of disruption/months |  | 12 |  |  |
| --- | --- | --- | --- | --- |
| Duration of background delay/months |  | 2 | 4 | 6 |
| No threshold | Cases detected | 11226 |  |  |
|  | Cases missed | 0 |  |  |
|  | Attributable deaths | 653 | 1419 | 2250 |
|  | Life years lost | 9214 | 20315 | 32799 |
|  | Colonoscopies required | 511394 |  |  |
| FIT>2<br>Sensitivity: 97% | Cases detected | 10889 |  |  |
|  | Cases missed | 337 |  |  |
|  | Attributable deaths | 20 | 43 | 68 |
|  | Deaths mitigated | 634 | 1377 | 2183 |
|  | Life years lost | 276 | 609 | 984 |
|  | Lost life years mitigated | 8938 | 19705 | 31815 |
|  | Colonoscopies required | 189216 |  |  |
| FIT>10<br>Sensitivity: 91% | Cases detected | 10216 |  |  |
|  | Cases missed | 1010 |  |  |
|  | Attributable deaths | 59 | 128 | 203 |
|  | Deaths mitigated | 594 | 1292 | 2048 |
|  | Life years lost | 829 | 1828 | 2952 |
|  | Lost life years mitigated | 8385 | 18486 | 29847 |
|  | Colonoscopies required | 97165 |  |  |
| FIT>150<br>Sensitivity: 71% | Cases detected | 7948 |  |  |
|  | Cases missed | 3278 |  |  |
|  | Attributable deaths | 191 | 414 | 657 |
|  | Deaths mitigated | 463 | 1005 | 1593 |
|  | Life years lost | 2691 | 5932 | 9577 |
|  | Lost life years mitigated | 6524 | 14383 | 23222 |
|  | Colonoscopies required | 38866 |  |  |

Since access to endoscopy is likely to be the tightest bottleneck in the CRC pathway, we modelled the consequences of implementing FIT to stratify access and reduce endoscopy dependency as a strategy to mitigate against delay in the care pathway. Adopting a FIT threshold of 10ug Hb/g faeces would reduce colonoscopy requirements by over 80%, while still identifying over 90% of CRCs in the symptomatic group. In the face of a 4 month delay, prioritisation of those with FIT >10ug Hb/g would mitigate 1,292 out of 1,419 predicted deaths per year and 18,486 out of 20,315 predicted life years lost. Adopting a higher FIT threshold of 150ug Hb/g further reduces colonoscopy requirements by an additional ~10%, but at the expense of mitigating only 1,005/1,419 of the deaths and 14,383/20,315 of the lost life years. Over the course of one year across England, using a FIT threshold of 10ug Hb/g rather than 150 ug Hb/g would save 287 lives and 4,103 life years but would require an additional 58,299 endoscopies (**Table 3**). A cut-off of 2ug Hb/g offers sensitivity of 97%, but only reduces colonoscopies to 37% of normal levels.

Addressing concern about COVID-19-related mortality in elderly patients in particular, we considered the risk/benefit trade-off of colonoscopy of a symptomatic patient. Even at very high rates of nosocomical infection (5% per procedure), for every age group in the symptomatic population, the benefit in cancer survival exceeded the risk of investigation. We next addressed the benefit of prompt investigation versus delaying a few months until risk of nosocomial infection rates could be expected to decrease. Below age 70, prompt colonoscopy essentially offers net survival benefit across the range of plausible rates of nosocomial infection. However, for those over 70, if the nosocomial risk is high (≥2·5% per procedure) then a delay is preferable if the risk of nosocomial infection declines during the interval (**Table 4**).

**Table 4:** Survival benefit from prompt colonoscopy versus delay for different rates of nosocomial infection (assuming no FIT triage).

| Nosocomial infection rate | Delay/ months | 30-39 years | 40-49 years | 50-59 years | 60-69 years | 70-79 years | 80+ years |
| --- | --- | --- | --- | --- | --- | --- | --- |
| 1% | 2 months | 0.09% | 0.09% | 0.08% | 0.06% | 0.04% | 0.01% |
|  | 4 months | 0.21% | 0.22% | 0.21% | 0.20% | 0.20% | 0.19% |
|  | 6 months | 0.35% | 0.38% | 0.36% | 0.36% | 0.37% | 0.36% |
| 2.5% | 2 months | 0.08% | 0.09% | 0.06% | 0.01% | -0.08% | -0.22% |
|  | 4 months | 0.20% | 0.22% | 0.19% | 0.14% | 0.07% | -0.05% |
|  | 6 months | 0.35% | 0.37% | 0.34% | 0.30% | 0.24% | 0.12% |
| 5% | 2 months | 0.08% | 0.08% | 0.03% | -0.09% | -0.29% | -0.60% |
|  | 4 months | 0.20% | 0.21% | 0.15% | 0.04% | -0.14% | -0.44% |
|  | 6 months | 0.34% | 0.36% | 0.30% | 0.20% | 0.02% | -0.28% |

Applying the same analysis for surgery following diagnosis with CRC, if rates of per-day nosocomial infection are very high (>10%), for those aged over 60 with Stage 1 tumours, a short delay is advantageous provided nosocomial infection rates can be expected to decline in that period (**Supplementary Table 2**).

## DISCUSSION

We quantify the impact on survival of delays to definitive surgery for CRC, taking into account mortality risk from nosocomial COVID-19 infection. We examine the impact on both overall survival and actuarial life years lost, with 10-year CRC survival approximating well to long-term survival. We present data for per-patient delays of two, four, and six months, and periods of disruption of one year and two years, but provide a flexible model by which the reader can explore other scenarios and the possibility to update the parameter values with newer evidence (*e.g.* on age-specific mortality from COVID-19 and procedure-specific nosocomial infection risk estimates) (**Supplementary Materials**). We present analysis of the impact of different periods of delay and variable durations of disruption, summed across the totality of CRCs diagnoses through the urgent symptomatic 2WW pathway in England. Predicting that availability of endoscopy represents the most significant bottle-neck on the pathway, we present analysis of FIT-triage of symptomatic patients. We demonstrate that FIT threshold of ≥10 ug Hb/g would reduce endoscopy requirement by 80% whilst still identifying >90% of CRCs. Risk-benefit analysis of diagnostic colonoscopy weighing CRC survival against mortality from COVID-19, shows net benefit of colonoscopy across all ages, providing the nosocomial infection rate at endoscopy is ≤1%. Risk-benefit analysis of surgery weighing CRC survival against mortality from COVID-19, shows benefit of prompt surgery (< 3 month delay) for daily nosocomial infection rate of ≤5%.

### Implications for patient management

Our analysis indicates clear benefit of prompt investigation of all patients with symptoms indicative of CRC in nearly all circumstances. If nosocomial infection rates are high (≥2·5%/procedure), for older patients (aged >70) delay is preferable to colonoscopy provided nosocomial rates will decline. Once diagnosed, risk-benefit generally favours surgery; for stage 1 tumours delay of 2 months in those aged over 70 is preferable to surgery in the face of high per day nosocomial infection rates (≥5%/day), again assuming nosocomial rates will decline.

### Implications for healthcare planning and resource utilisation

Delay to diagnosis results in tumours being more advanced, resulting not only in poorer survival, but requirement for more costly surgery and/or chemotherapy to manage the upstaged disease. Our per day hazard ratios include the reduced survival for the appreciable number of CRC patients who will present during the period of delay as emergencies (e.g. bowel obstruction, perforation, or bleed). However, resource requirements will also be dramatically higher for these patients, for example use of emergency theatre and ICU stay^21–23^. Due to the combination of progression and increased likelihood of emergency presentation, even modest delays to diagnosis of CRC will have significant impact on mortality and life years lost.

As lockdown is lifted, there will be a ‘bulge’ of symptomatic patients presenting to primary care, which will need to be addressed alongside processing of the normal stream of incident cancer presentations. Unless supranormal capacity is made available within each of primary care cancer, diagnostics, and surgery ahead of ease of lock-down, knock-on per-patient delays may persist for months or even years. Given chronically limited capacity, endoscopy services are likely to be under the most acute and prolonged stress. Urgent review is required of British Society of Gastroenterology guidance, to clarify their position on colonoscopy for symptomatic patients, with development of improved protocols protecting staff undertaking these procedures^5^. Although not currently standard practice for symptomatic patients, FIT screening confers the opportunity to rationalise limited endoscopy resource. It is however desirable that FIT thresholds be much lower than those used in population screening programmes or a significant proportion of CRC will be missed.

It is essential in the medium-long term, beyond recovery from the first bulge, that pathways for CRC referral, diagnostics, and surgery are restored to normal capacity and function without excess delay. Whilst FIT-screening at a threshold appropriate to a symptomatic population provides a rational means for prioritising cases; patients ‘negative’ at that threshold with ongoing symptoms will nevertheless ultimately require investigation.

As our analysis demonstrates, for those patients aged s >70 years in which CRC is common, the per case benefit of colonoscopy and surgery are predicated on the likelihood of nosocomial infection. Rigorous protocols for staff screening and shielding are required, along with active focus to establish ‘cold’ sections of the healthcare system for both diagnostics and surgery. This will not only serve to reduce mortality from nosocomial infection, but will also provide reassurance to the public regarding uptake of diagnostics and surgery for cancer.

### International relevance

While we have used data for England, CRC diagnosis, management, and survival are directly comparable across most economically developed countries so the impact of delay is broadly applicable. Our model will provide insights highly relevant to other countries and health system settings, provided that the population structure and background rates of population mortality unrelated to colorectal cancer causes are similar to those of England. We would like to highlight that our model only reflects urgent elective diagnosis and management; there may be international variation in proportions of CRC cases presenting through the different routes (urgent symptomatic, screening, emergency, routine).

### Limitations of analysis

The accuracy of our predictions is predicated on the validity of assumptions and estimates used for parameterisation, as for any model-based analysis. Our approach is solely survival-focused: a more elaborate model capturing stage-transition may offer additional utility for healthcare planning. We have not considered the possibility of complex, serial, or interdependent bottle-necks, instead modelling a simple total delay to treatment. We have assumed that all urgent 2WW colorectal referrals from primary care result in colonoscopy; data are not available to specify the proportion diverted in secondary care to alternative investigation. In total 0·6% of 2WW colorectal referrals lead to the eventual diagnosis of another cancer type^8^. We have not captured the impact of delay on diagnoses of these cancers, nor how a FIT-based triage would impact on this.

We have not evaluated the impact of any changes in systemic anti-cancer therapy (SACT), bearing in mind that SACTs makes comparatively limited contribution to CRC survival, in particular for rectal cancer. As the primary focus of our analysis is delay to surgery with curative intent, we have not modelled impact of delay on stage 4 outcomes, instead making the assumption that treatment delay has no impact on mortality (*i.e*. that any survival benefit from SACT management of stage 4 disease would be counterbalanced by risk of COVID-related mortality).

We have also focused exclusively on detection of invasive CRC. Due to lack of data on the frequency and distribution of pre-invasive adenomas along with robust observation data by which to model progression, we have not be able evaluate the impact of detection and removal of non-invasive adenomas in our analyses. Hence, our estimates of the impact of delays in care provision will be inherently conservative.

We have focused exclusively on pathways for symptomatic patients and have not considered CRC ascertained via screening and ‘routine’ referral. The ‘routine’ route to diagnosis includes both patients referred non-urgently for symptoms and those under long-term surveillance for polyps, high risk conditions and/or family history. Whilst our analyses of tumour progression would be applicable in both contexts, impact and mitigation of disruption to these routes require specific consideration.

### Further research

In our current model, we estimate the effects of a specified period of per-patient delay. Dynamic models are required to enable the prediction on outcomes of (i) differential prioritisation and deprioritisation of patient groups, (ii) different patterns of re-presentation of ‘accumulated’ cases alongside incident cases, and (iii) varying release of bottlenecks in primary care, diagnostics, and surgery. Given competition for specific groups of healthcare workers, resource-focused and health-economic analyses would also be of value. We have focused on the impact to surgery with curative intent; analyses are also required to quantify the impact on mortality of withholding of life-extending chemo-and radio-therapy for patients with Stage 4 disease.

## CONCLUSION

Delay to treatment for CRC results in significant mortality and lost life years. Providing nosocomial infection rates are not high, diagnostic colonoscopy and surgery with curative intent offer survival benefit in all age-groups. Prioritisation of symptomatic FIT-positive patients is a rational approach to prevent delay if access to endoscopy is limited.

## Data Availability

This work uses data that has been provided by patients and collected by the NHS as part of their care and support. The data are collated, maintained and quality assured by the National Cancer Registration and Analysis Service, which is part of Public Health England (PHE).

## AUTHOR CONTRIBUTIONS

C.T., M.E.J., A.S. and R.S.H. designed the model. M.E.J. provided cancer progression models. J.B. generated and quality-assured the NCRAS datasets applied to the model. M.A. and C.B. provided data on FIT performance from the NICE FIT study. M.E.J., J.B. C.T., R.S.H., A.S., E.R. M.L., E.M., G.L., D.C.M and M.W., provided epidemiological expertise in parameterisation of the model and appropriate literature. S.A.B, S.J., D.L.N, E.W and M.W. provided details of clinical pathways and estimates of clinical resourcing. B.T., A.G. and C.L. quality assured and user-tested the model. C.L and B.T. assembled figures for presentation. C.T drafted the manuscript, with substantial contribution from A.S., R.S.H., M.E.J., G.L. and C.L.. All authors contributed to the final manuscript.

## ACKNOWLEDGEMENTS

MEJ additionally received funding from Breast Cancer Now. B.T and A.G. are supported by Cancer Research UK award C61296/A27223. C.L. and C.T. receive support from the Movember foundation. R.S.H. is supported by Cancer Research UK (C1298/A8362) and Bobby Moore Fund for Cancer. GL is supported by a Cancer Research UK Advanced Clinician Scientist Fellowship Award [C18081/A18180] and is Associate Director of the multi-institutional CanTest Collaborative funded by Cancer Research UK [C8640/A23385]. Research UK). D.C.M is supported by Cancer Research UK (C57955/A24390. A.S. is in receipt of an Academic Clinical Lectureship from National Institute for Health Research (NIHR) and Biomedical Research Centre (BRC) post-doctoral support. This is a summary of independent research supported by the NIHR BRC at the Royal Marsden NHS Foundation Trust and Institute of Cancer Research. The views expressed are those of the authors and not necessarily those of the NHS, NIHR, or the Department of Health. This work uses data that has been provided by patients and collected by the NHS as part of their care and support. The data are collated, maintained and quality assured by the National Cancer Registration and Analysis Service, which is part of Public Health England (PHE).

## RESEARCH IN CONTEXT

### Evidence before this study

Coronavirus disease 2019 (COVID-19) has resulted in disruption across cancer diagnostics and surgery, affecting colorectal cancer (CRC) in particular through widespread shutdown of routine endoscopy due to safety concerns and under-staffing. Observational studies have quantified the impact on long-term survival of delay to treatment in CRC, albeit with inherent confounding by indication. Routinely generated data from PHE demonstrate varying proportions between tumour type presenting via urgent symptomatic, routine, screening and emergency routes. There has been no systematic evaluation of the impact on survival of global delays to diagnosis focusing on specific routes. Most data on FIT pertain to application for screening; only small studies of FIT in symptomatic patients.

### Added value of this study

To our knowledge, we provided the first explicit analysis of the impact on long-term survival of delay to CRC diagnosis in symptomatic patients. Combining detailed data on cancer surgery in England 2007–2017 from the National Cancer Registration Analysis Service with per-day hazard ratios of CRC progression from observational studies, we quantified the impact of different periods of per-patient delay on CRC survival in symptomatic patients, both for age-specific stage-specific individual tumour groups and summed for annual case volumes. We provide the first explicit analysis of FIT triage to mitigate impact, examining thresholds of 2, 10, and 150 ug Hb/g. We provide the first explicit analysis of the impact of different rates of nosocomial COVID-19 infection on survival benefit of diagnostic endoscopy.

### Implications of all the available evidence

Colorectal cancer is common with high mortality. Delays in urgent referral of symptomatic patients due to bottle-necks in endoscopy has the potential to cause high attributable deaths and lost life years. FIT triage at 10 ug Hb/g offers opportunity to mitigate >90% of these deaths and reduce exposure of patients to nosocomial COVID-19 infection.

